# Informal social support for chronic pain and distress in people with HIV: identifying targets for intervention

**DOI:** 10.1101/2025.11.07.25339650

**Authors:** Victoria J Madden, Lara Court, Andiswa Gidana, Lucia Knight, Antonia Wadley, Luyanduthando Mqadi, Stephan Rabie, Robert R Edwards, John A Joska, Romy Parker

## Abstract

Social factors are consequences and determinants of chronic pain, yet little is known about how informal social support operates for people with chronic pain and HIV. The overlapping burdens of emotional distress and chronic pain in people living with HIV raise the need to identify targets for culturally resonant treatments. This qualitative study explored how people living with chronic or recurrent pain and HIV seek informal social support for pain and distress from their social networks, how these networks operate, and how social support is influenced by socio-ecological factors. Adults with virally suppressed HIV who repeatedly reported pain were purposively sampled and interviewed. Semi-structured, individual in-depth interviews (n=18) explored individuals’ experiences of living with HIV and chronic pain, distress and its relationship to pain, and help-seeking behaviours for chronic pain and distress. Codebook thematic analysis was used within an interpretivist epistemological approach. Support for pain involved more tangible actions by fewer people than support for distress. Disruptions in social support were more likely when the person with pain was irritable, socially withdrawn, or unable to fulfil social roles. Interpersonal conflict could also cause or increase pain and distress. Current pain assessments rarely identify the social interference of pain identified here. This work identified potential targets for individual-level interventions to sustain informal social support: individuals’ abilities to cope with daily stressors, pain self-management strategies to facilitate socialisation and engagement in valued life activities, and strategies to prevent support fatigue in informal social networks.

## Introduction

Chronic pain is the world’s most burdensome health condition in terms of years lived with disability (1, 2). People with chronic pain incur significant difficulties, such as comorbid health problems, limited ability to work, and high healthcare costs (3).

Chronic pain has far-reaching impacts, extending beyond the individual with pain to their families, communities, and societies. It adds financial and emotional strain to the family unit as a whole (4). It increases the risk of chronic pain in younger generations, especially if a parent has chronic pain and comorbid depression (5–7). Community members with disabling pain make smaller monetary and practical contributions, while drawing on more resources than their pain-free peers, thus compromising reciprocated support (4, 8). A high prevalence of health problems in a single community could thus deplete both tangible and emotional community resources, particularly in contexts where social security systems are inadequate or absent—leaving people with pain, and their families, relatively isolated.

Social factors are recognised as both consequences and determinants of chronic pain. For example, chronic pain can introduce conflict and relational dissatisfaction, along with guilt for the person with pain and their family and friends – and lower quality of relationships in the long term (9–11). The interference of pain in work and community activities progressively isolates those with pain, contributing to loss of roles and sense of identity (12). Isolation or loneliness can compromise immune function (13, 14), increase risk of mental illness (15), and constrain cognitive function that is needed to live well with chronic health conditions (16).

Conversely, social support can improve health in people with chronic pain, partly by buffering pain-related emotional distress and facilitating adaptive coping, and possibly also by improving regulation of physiological responses to painful events (17, 18). Pain-related emotional distress is known to exacerbate pain and mental concerns, which gives the distress-relieving impact of social support relevance to both chronic pain and mental health (19). The health benefits of social support are highest when individuals perceive their social network to be available and supportive (20, 21), which is shaped by their experiences receiving support when they need it, and by the impact of the support (22). However, it is crucial to recognise that the interpersonal relationships and social networks that underpin social support are inherently dynamic, so the social support available to an individual may change over time.

Despite the ripple effects of chronic pain and the centrality of social support, little is known about how social support influences, or is influenced by, chronic pain in the context of HIV (23–25). This is surprising in light of the high burden of chronic pain in people with HIV (26–28). It also contrasts starkly with both the HIV treatment adherence literature—where social support interventions are already being used to improve treatment adherence—and with a consensus-based agenda for research on HIV-associated pain that included social support as a prioritised study outcome (29, 30).

In South Africa, the dual burden of HIV and chronic pain is well positioned to compromise population health: the national prevalence of HIV is 20%, and, similarly, 20% of South Africans reports chronic pain (31). Recent qualitative research noted that rural South Africans with HIV varied in how they disclosed pain and that disclosing pain opened opportunities to receive interpersonal support (32). However, little is known about how social networks function to address pain and emotional distress in the context of chronic pain and HIV in South Africa. Given the constrained resources in the South African healthcare system and that social factors may be modifiable determinants of health that could be targeted by scalable, culturally relevant interventions, this study aimed to explore (1) how people living with chronic or recurrent pain and HIV seek informal social support to alleviate pain and emotional distress, (2) how informal social support networks function, and (3) how socio-ecological factors influence informal social support, in the context of chronic or recurrent pain and HIV. We hoped to identify opportunities to develop and test individual-level or community-level interventions to leverage and strengthen existing informal social support systems, to improve health among people with HIV and pain. Here, informal social support is differentiated from support derived from formal professional, or public services (i.e. health clinic, social welfare services) (33), and understood as being received through informal relationships within an interactive field of people – a social network (34, 35). Social network *functions* relate to the *received* and *perceived* social support provided by social network members, which can include emotional, tangible, and informational support, as well as companionship (Table 1) (36). In this work, emotional distress refers to negative affective states such as worry, anxiety, depression (19), and includes feelings attributed to pain or to other difficult circumstances and experiences. The closest isiXhosa term for this draws on a metaphor of ‘the weight you’re carrying on your shoulders’ and is interpreted as describing psychological pressure or strain. The philosophical perspective for this work was an interpretivist epistemological framework; we sought to understand the knowledge and meaning that our participants had attached to their experiences of seeking and receiving informal social support for this distress and pain, informed by their social, cultural, and broader context.

**Table 1:** Social support theory constructs relevant to this study (37–39).

| Types of social support | Definition |
| --- | --- |
| Emotional support | Provision of listening support, provision of emotional comfort and security, increasing the person's perceived sense of competence. |
| Tangible support (also known as instrumental support) | Provision of concrete assistance, such as giving time, providing transportation, or giving financial assistance. |
| Informational support | Giving of advice, suggestions and guidance; sharing of information. |
| Companionship | Involves the sharing of leisure and other activities motivated by the internal goal of enjoyment and wellbeing; understood as both a relationship quality and |
|  | a form of social support. |

## Methods

### Study setting

This exploratory qualitative study is an extension of a ‘parent’ quantitative longitudinal study of 99 people living with virally suppressed HIV and chronic pain or no clinical pain (40, 41), serviced by standard HIV care (including free antiretroviral treatment, ART) at a primary health facility in a peri-urban community near Cape Town, South Africa. The parent and current studies were approved by the Human Research Ethics Committees at the University of Cape Town and City of Cape Town (approvals 764/2019 and 24699). The parent study focused on quantitative relationships between distress, inflammatory responses, and pain, by using questionnaires, blood tests, and psychophysical testing over an initial six months, during which participants were also invited to respond remotely to weekly questions on their pain and distress for up to six months. For the current, qualitative study, participants had to have repeatedly reported pain. Therefore, we purposively selected participants who had repeatedly reported pain in those quantitative data and recruited them in two waves, prioritising those reporting pain either consistently or repeatedly (a sampling frame of n=42). The main data collector for the study (AG) telephonically invited participants to an in-person, individual interview (n=17 could not be contacted; n=2 declined; n=23 agreed to interview). The final sample of 23 was determined by participant availability. In-depth interviews were completed by 18 participants (n=5 did not arrive), which was deemed appropriate to contain adequate information power for the topic (42). All participants gave additional written informed consent. In this peri-urban community, approximately half the people live in informal housing, one in four is unemployed (43), and the density of HIV-positive people is thought to be the highest in the province, with HIV prevalence between 10 and 22% (44, 45).

### Data collection

The data presented here were drawn from these individual, semi-structured in-depth interviews conducted between 20 June and 26 August 2022, in a private room adjacent to the health facility, to capture rich descriptions of inter-individual differences and intra-individual variations in behaviour and experiences in a confidential manner. Topics explored individuals’ experiences of living with HIV and chronic pain and perceptions of how these affected their lives; experiences of distress and its relationship to their chronic pain; coping strategies and help-seeking; formal healthcare and formal and informal social support, and behaviours used for chronic pain and distress. The interview guide was consultatively developed, based on the topic of the larger study, existing literature, and identified gaps relevant to the local setting according to key expert and local knowledge (JAJ, AG, AW). Interviews were conducted in participants’ preferred language (i.e. either English or IsiXhosa), by a trained and experienced female bilingual isiXhosa-English interviewer (AG), who was resident in a neighbouring community. The interviewer already had established, confidential relationships with all participants and had previously asked personal questions of them (i.e. about mental health, income, social support, stressful life events and traumatic childhood events). Participants provided written informed consent. Interviews took 56-122 minutes, during which participants were offered refreshments. The interview period coincided with the COVID-19 pandemic restrictions and major electricity cuts in South Africa, resulting in minimal light or static torchlight for some interviews. Participants were compensated for their time with R150 (∼US$9). Interviews were audio-recorded. The isiXhosa interview recordings were later verbally translated into English audio files by AG, with an explicit priority to preserve semantic equivalence and contextual meaning. The English audio files were then transcribed using the online, HIPPAA-compliant OTTER software (https://otter.ai). All audio files were retained. The transcriptions were independently checked against the English audio files by one of two research assistants, to correct errors in transcription. English words that were difficult to discern or that seemed not to make sense in context were checked against the isiXhosa recordings and, when necessary, discussed with AG. Words and sounds that remained unclear were marked as such in the transcripts. The checked transcriptions then moved forward to analysis.

### Data analysis

Codebook thematic analysis was used (led by LC) (46, 47). Following familiarization, transcripts were coded in qualitative data analysis software, NVivo 15 (Lumivero, London, United Kingdom). An initial codebook was developed according to primary study objectives and interview guide sub-sections (i.e. lived experiences of HIV and chronic pain; functional impacts of HIV, chronic pain and distress; help-seeking strategies for pain and distress) (48), which was then shaped further by initial inductive coding of four transcripts. To remain consistent with the relevant body of knowledge (34, 49, 50), themes and subthemes identified using the initial codebook were refined, recategorized, and named according to social support theory constructs, where appropriate, specifically to describe the structure and function of social support (see 2.2.a and 2.2.b) (51). Inductively developed codes were used to generate themes regarding socio-cultural factors influencing the structure and function of social networks (see 2.2.c). AG reviewed data interpretations. Consistent with an interpretivist epistemological approach, results and processes were iteratively discussed (LC, LK, VJM) to facilitate meaning-making and reflexivity and an audit trail was kept, using journals, memos, and notes (52, 53). The Consolidated Criteria for Reporting Qualitative Research (54) was used to guide reporting, omitting identifying data. The lead and support analysts are all South African women. LC (Occupational Therapist, MPH) and LK (PhD) have led qualitative research on health experiences of South Africans and had no contact with participants; VJM (Physiotherapist, PhD) is a clinician-scientist focused on pain who led the parent and current study and exchanged transient greetings with two parent study participants; AG (PGDip) is a Xhosa woman research assistant, was the primary in-person data collector for most of the parent study, conducted the current interviews, and reviewed our data interpretations for congruity with the interviews to counteract any potential misinterpretation by non-Xhosa women LC, VJM, and LK.

## Results

### 1. Participant characteristics

Interviews were completed with 18 participants; 17 in isiXhosa and 1 in English. Four participants lived alone and the median (IQR) monthly household income was R1845 (R1225-3825), which is 23% of the national median and 13% of the City of Cape Town median reported for late 2022 (55).

### 2. Findings

First, we describe the *structure* of social networks that provided informal received social support to participants to alleviate pain and distress. Second, we describe the *functions* of social networks, specifically how members provided emotional, informational and tangible social support and companionship for a) chronic pain and b) emotional distress, together with the perceived impact of this support. Third, we explore the socio-ecological factors influencing the structure and function of social networks in the context of chronic pain, presented under themes of how 1) chronic pain causes emotional distress, resulting in social isolation, 2) chronic pain affects one’s ability to perform social roles, and 3) inter-personal relationships can cause emotional distress and pain. In line with an interpretivist epistemological stance, all findings presented here rest on the subjective perspectives of the research participants and should not be conflated with objective causal evidence.

#### 2.1. Structure of social support networks

Participants described seeking and receiving informal social support for pain and emotional distress from a range of family and community members, which are displayed in Figure 1. Support for chronic pain came from a smaller social network, predominantly partners and immediate and extended family members (i.e., children, parents, sisters, aunts, and cousins). Others included people in their ART adherence club at their local clinic (into which people are assigned when they first achieve viral suppression), and close confidants, such as friends and neighbours. When in distress, participants reached out to these members as well as an additional, wider group of social network members, including church leaders and fellow churchgoers, work colleagues, and people they sat next to at their local clinic or on public transport.

**Fig 1:**
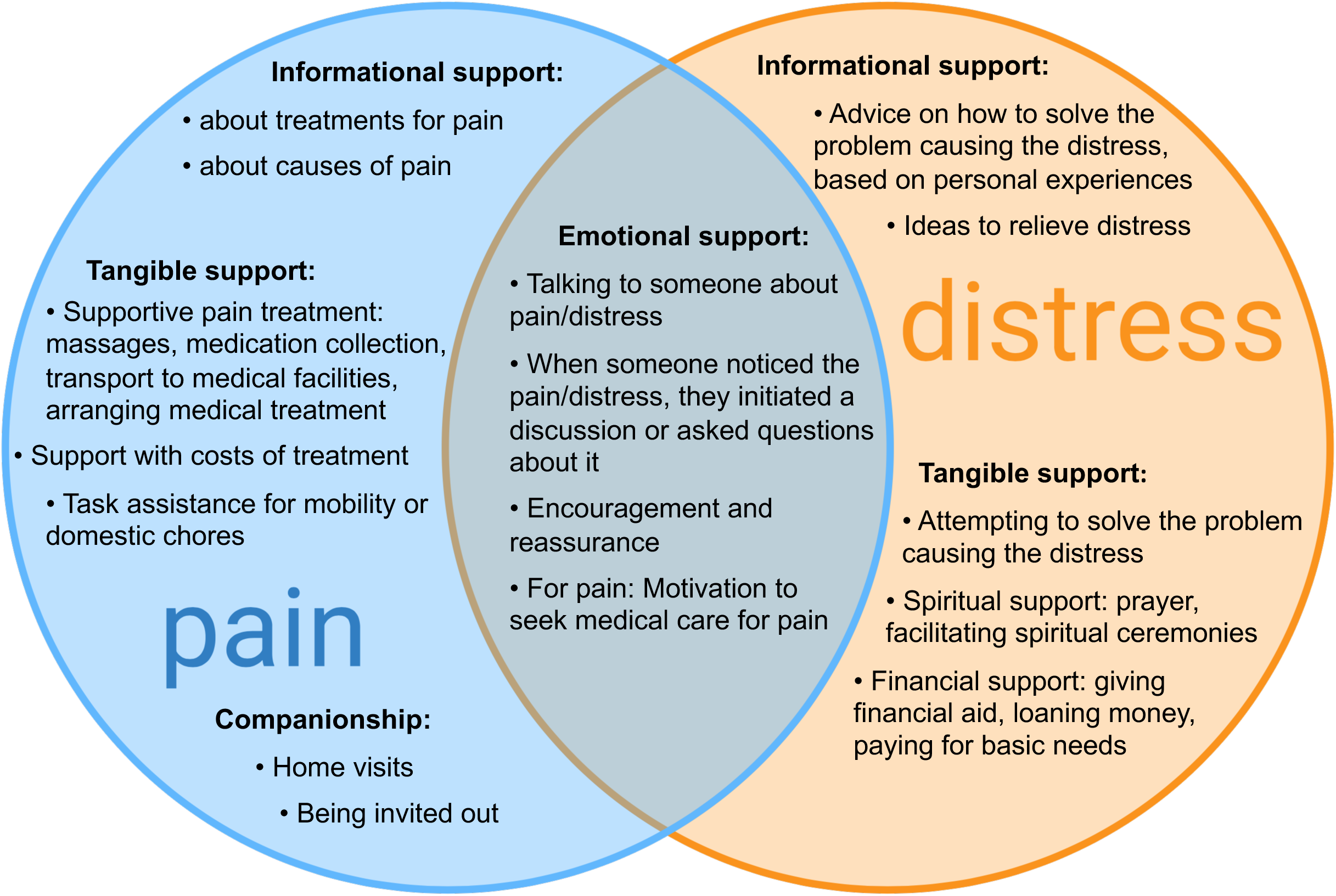
Visual depiction of members of social network accessed for social support for both chronic pain and distress (blue & orange) or distress only (orange)

#### 2.2. Functions of social support networks

Participants received a range of informal social support from social network members, discussed below under categories of emotional, informational and tangible support functions, together with the perceived impact of this support. Figure 2 provides a visual thematic summary of the overlapping and distinct support functions for pain and emotional distress, described in detail below.

**Fig 2:**
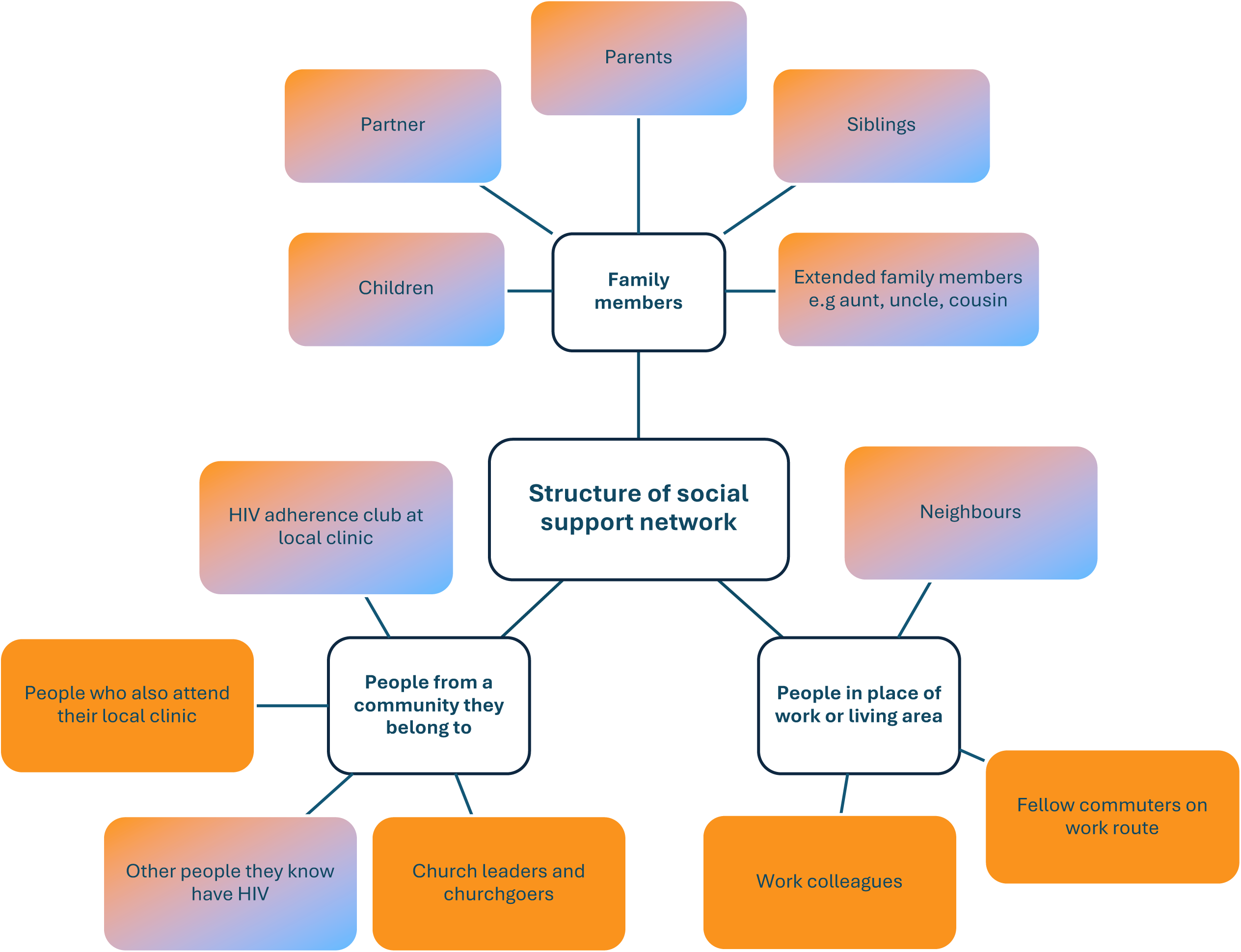
Informal social support received from social network members for pain and emotional distress, indicating unique and overlapping features.

##### a. Received informal social support for chronic pain

Participants framed disclosure of their chronic pain to social network members as a way of seeking emotional support. Disclosing their chronic pain was challenging in instances when they did not want others to know about their pain, as it might worry or burden them, or if they were not talkative in nature. Participants preferred to disclose their pain to people they lived with or saw often, and close confidants, who were “always there for them” and willing to listen. Some of these trusted social network members were people they had previously disclosed their HIV status to, or others they knew were HIV positive themselves, as they felt they could “tell them everything”:

> *There is someone who I would tell when I’ve got the pains… so I’ll go to this a neighbour of mine, the one that I told you that is also HIV positive, I will go to her and then I will talk with her… about the things that I’m feeling. (female in 50s, living with partner and children)*

Conversely, members who were perceived as responding negatively to hearing about their initial HIV diagnosis were not sought out for support for pain. Participants felt cared for and understood when social network members noticed, asked about, or initiated discussions about their pain. This solicitous emotional support typically came from those who lived with or near them and saw them in pain, or from close confidants who phoned them. Participants who lived on their own described feeling supported when social network members provided solicitous support in conjunction with companionship, by visiting them, or by inviting them to visit:

> *Iwould tell my aunt [about pain], I’ll just call her and tell her that I’ve got the pains… She [aunt] will come and visit me and see how am I doing… [and] I become fine,… so, if she comes, at least that makes a difference. (female in 30s, living alone)*

Provision of encouragement also eased pain-related emotional distress. This was experienced when members motivated participants to seek formal medical care for their pain, reassured them, or encouraged them not worry about their pain:

> *You know, my daughter is always there for me, and she’ll tell me that, “No, Mama, you are going to be alright you know”. (female in 30s, living with partner and children)*

Although demonstrative support in response to disclosure of their pain was appreciated, the process of “sharing” their pain was what helped some participants feel “happy” and hopeful. Talking to someone about their pain was often enough to relieve pain that the participant attributed to emotional distress (a common attribution for headaches), or to give them confidence about managing their pain:

> *I do tell my cousin, my family, they know when I’ve got the pains, I will tell them. Everyone. Even with my mother, I’ll phone her and tell her that I’ve got these pains… I’m just happy that I have talked… (female in 30s, living with family)*

In addition to motivating participants to seek formal medical care, members would often provide informational support. Pain-focused informational support was often sought from members who also had chronic pain or friends who were health professionals, as they were felt to be able to provide “good” advice. Members would advise on what could be causing the pain, based on their own experiences or personal knowledge:

> *I will tell my neighbour that I’ve got this pain,… they’re the one that advised me to go buy pills for arthritis… (female in 40s, living with children)*

Members also advised on which health provider to see and recommended different medicinal or physical treatments for pain. Participants explained that they appreciated the advice, even though it didn’t always relieve the pain:

> *… then they will advise me that, okay, I must use this must use [or] that.… I tried it, but it didn’t help me. But I’m happy with the support that I’m getting from them. (female in 40s, living with children)*

Tangible support received was aimed at supporting treatment of pain or assistance with tasks that participants found challenging due to pain. Participants described how family members, both nuclear and extended, sent money or paid for their direct or indirect medical costs, drove them to and arranged care for them at health facilities, fetched their medication from the health facility, or shared their own medication with them. Family members living with them commonly provided massages using rubbing ointments or wrapping materials:

> *It’s only my kids that knows that I have got pains… The one…will rub me, massage my legs, wrap my neck… they go buy pills… or massage me. (female in 40s, living with family)*

Although some participants with less disabling chronic pain described not needing help with their tasks and preferred to “force” themselves to do tasks, assistance with tasks that were challenging due to pain was described by many as being supportive. This was predominantly provided by members living in their household or those who lived nearby and were able to provide physical support:

> *He [partner] will get me water and do the washing because now I can’t go to the tap, the tap is far and I can’t carry heavy stuff… I have to get the kids to go and get water for me… (female in 50s, living alone)*

Support included assistance with getting out of bed or moving around the house, bringing their medication to them, or performing their domestic tasks, such as helping participants to cook and prepare meals, wash clothes or dishes, or clean their house.

##### b. Received informal social support for emotional distress

Participants described a range of socio-ecological factors, particularly unemployment and a lack of financial resources, as contributing to stress, anger, depression, and anxiety. Some who were employed described challenges with their employment, such as conflicts with employers (i.e. who did not appreciate their work, treat them well or follow fair labour practices), living far from their workplace, or being unsatisfied with their type of work and salary. Although many received social grants, the grants did not cover the family’s basic needs, such as food, electricity or cooking gas and transportation, which caused significant daily emotional distress. Insufficient finances also meant that they could not realise personal goals and lived in environmental conditions that also worsened their pain:

> *When it’s raining, the water will get into the house… and there is no electricity there…I feel bad…at night the cold really affects me… I’ll have these chest pains… But I’m staying there because I don’t have any other options. (female in 50s, living alone)*

Participants described how talking to someone about their emotional distress helped them find relief and resulted in them feeling “lighter”, as they had shared their problems with someone else. This could be true even if a member did nothing more than listen. Others described how they felt emotionally supported when members encouraged them not to worry, especially if the reasons for their distress were out of their control, and when a member gave them hope by telling them that “things will be alright”, or expressed a belief that they had the capacity to cope with their problem. They also felt supported when others checked in on them and provided space to talk about their stressors:

> *The person that I talk to when I’m stressing is my partner… she’s the one who will notice that I’m not my usual self. She’ll ask me [about] what stresses me, then she will tell me that I need not stress a lot because there’s nothing that I can change about this situation. (male in 40s, living with partner)*

Emotional support for distress was sought from a variety of social network members depending on their availability and whether it was appropriate to seek their support. Available members were those who were seen frequently at work or on public transport, or who made themselves available to talk to when needed. A few participants specifically chose to talk to “strangers” regarding commonly experienced stressors or due to their being external to the distressing situation:

> *I’ll talk about my stuff with strangers. I know those people don’t know me. But… if I can just speak to a stranger, someone who doesn’t care about me - I feel relieved about that. (female in 30s, living with family)*

In contrast, others expressed that it was not appropriate to share their emotional distress with “just anyone” as these people might reveal their problems to others or “make fun” of them. These participants preferred to choose members they felt comfortable with and had an open and trusting relationship with, who listened, would not judge them. Being empathetic to their distress was a key consideration:

> *I do talk to my sister that I’m staying with, but not everything with her. Because she is not that kind of a person who listens or who is interested. You could see, even when you’re telling her something, she doesn’t respond, you know? (female in 30s, living with family)*

Participants also described valuing advice on how to deal with their stressors. Informational support was provided by members sharing their own experiences with a similar problem and offering advice born of these experiences, often including strategies to solve the problem or how to cope with the resulting emotional distress. This was described as relieving participants’ distress to a degree:

> *I will speak to the ladies that I go with… there [at the clinic]… I will normally speak to them, like try to find out… how do they cope with certain things in life, then I’ll get advice from them… we’ll share our experiences and problems… it is good to share your problems with other people. (female in 40s, living with family)*

This support was often sought from members who knew about the participant’s stressors or if they had perceived experience with the same stressors. For example, family members who knew their family dynamics were sought out for advice in times of sibling conflict, or members with similar-age children were engaged with for informational support on how to manage their own children’s distressing behaviours:

> *I do get the support from my partner because she’s the one that I always talk to when I’ve got these things that are stressing me…her child is [a young adult], so she’s been in the situation that I’m in. She knows how to deal with the teenagers and then she’ll advise me. (male in 40s, living with partner)*

Tangible support was provided in the forms of financial support, spiritual support, or helping to solve the problem that was causing the emotional distress. Participants described how, after talking about their emotional distress, members offered to do something to solve the problem, such as brainstorming business ideas with them, talking to their “troublesome” child, or handing in the participant’s CV to employers. Spiritual support was also valued. Participants described discussing their stressors with church members who would pray with them, and ceremonies facilitated by spiritual leaders also helped to relieve their distress:

> *It has given me strength, ever since I started to go into that church, in terms of the way that I think, it has changed me…So when I prayed to those candles there, so it’s helping me spiritually. So, they’re [spiritual leaders] using candles and coarse salt, you know, salt water. It’s normal water that we’re using, you just have to bring water [to church] and then they have to pray over that water, and then they will maybe do what we call ‘isiwasho’ [salt and water], and then you drink that ‘isiwasho’… bath yourself [with it]… you can spray [it] around your house for bad spirits. (female in 50s, living with family and children)*

Economic support included giving or loaning money, sometimes for specific purposes, such as to support a particular household member or pay for basic needs.

#### 2.3. Factors influencing the structure and function of social networks in the context of chronic pain

Participants described how their social support structure and functions were influenced by their chronic pain. They described a) how chronic pain caused emotional distress, resulting in isolation; b) how pain affected their ability to perform social roles, and c) how interpersonal relationships could be both a prominent source of informal social support and a cause of emotional distress and pain.

##### a. Chronic pain results in emotional distress, leading to social isolation

Chronic pain impacted on participants’ mood and emotions and made them want to isolate themselves from others. Notably, although some mild emotional distress was experienced due to HIV (i.e. due to stigma experienced from health care workers, when a partner died from HIV, or due to the need to take medication daily), many of our participants described how their HIV did not cause emotional distress beyond the initial period of diagnosis and initiating treatment, yet their chronic pain often resulted in ongoing distress:

> *The other things that are stressing me about my health, are the pains that I have. They are the most thing that are stressing me. HIV doesn’t stress me, no. The fact that I’m on ARVs [antiretroviral treatment] doesn’t really stress me. (female in 50s, living with partner and children)*

Participants described experiencing negative emotional responses to their pain, which continued even in periods when they experienced no or mild pain, as they felt anxious that the pain could recur at any time. Overarching low moods were described, such as feeling “irritable,” “bad” and “not happy”, as well as frustrated, angry and stressed. These were due to the pain’s chronic nature, a lack of diagnostic certainty, few pain-relieving strategies, and the current and possible future impacts of their chronic pain, specifically on their mobility, independence and ability to work:

> *Sometimes I’ll get afraid of the pains, when I don’t know what is happening and I’ll be wondering why it doesn’t go away… now I don’t know which pain tablets should I take, because they don’t necessarily work…I’m afraid that I might not be able to walk… I don’t know what is going to happen in future… I feel bad. (female in 50s, living with family)*

Experiencing chronic pain also brought with it additional fears. For example, those with diagnostic uncertainty expressed fear of death, as they were unsure if their chronic pain was related to something terminal. Chronic pain was also related to aging, leading some participants to describe a fear of getting old:

> *Now you’ve got that fear that oh, now I’m getting old and growing, I am going to have pains like my mother because my mother is always like, crying about having pains. (male in 30s, living with partner and children)*

Emotional distress resulting from pain influenced participants’ interpersonal relationships with those in their social network. Feeling irritable due to their pain was described as affecting their relationships with those they lived with, who grew tired of their complaints about pain:

> *I’ll be complaining that I’ve got the pain here, I’ve got the pain there, you know… I’m getting irritable all the time. Now [my husband]’s been saying that… I’m always complaining about the pains. So… it was interfering with the relationship…with my kids and my husband. (female in 20s, living with partner and children)*

Participants commonly described wanting to be alone when feeling emotional distress due to their pain or when experiencing pain. They described intentionally isolating themselves at home by withdrawing to their rooms or avoiding seeing friends and other family by staying at home more often, particularly when experiencing pain. They also described ignoring phone calls, not speaking to others at social gatherings, shortening their social visits with others, or cancelling them altogether:

> *Sometimes you find that it [the pain] will make me not be able to go,… I’ll feel like I don’t want to go anywhere… just want to stay at home.…I’ll end up not going or cancelling the things that I was supposed to do. (female in 30s, living with family and children)*

Participants described varied motivations for isolating themselves, typically feeling disinclined to talk to others when in pain, wanting their “own space”, feeling tired or “lazy” to go out, or wanting to be somewhere comfortable when in pain. Participants also avoided social gatherings because they did not want others to worry about—or in other cases, not know about—their pain:

> *When that [pain] happens, I just want to be alone. I don’t want to be seen by other people because… I don’t want them to know about my problem. (male in 40s, living alone)*

Participants described how isolating themselves affected their social relationships. Although some friends and family continued to visit them at their home when they stayed at home more often, they often started “meeting” people less often and neighbours and friends were described as complaining that the participant had “been scarce”. This meant that fewer people knew about their pain and could offer support.

##### b. Chronic pain affects one’s ability to perform social roles

Participants described how chronic pain affected their ability to perform role-specific tasks, influencing the functions of their social network. This was contrasted to the daily impact of HIV. Limited challenges of HIV were noted, such as negotiating intimate relationships with an HIV positive status or engaging with family or friends while anticipating stigma. The impact of HIV on social roles, such as parenting, working, and community roles, was felt to be minimal and some participants even noted improvements in support:

> *My [social] responsibilities have never changed as much [compared to pain]… So, when I told them [my family] about my status, the only thing that we discussed was the way forward… So ever since then, my support system has been alright..*.
>
> *Otherwise, I don’t see any difference with my life now… even with my [intimate] relationship, I’m aware of everything that I have to do and I’m learning new things. (male in 40s, living with partner)*

In contrast, participants described how chronic pain reduced their ability to provide certain tangible support for their children, perceived as crucial to the role of being a parent. Such activities included attending sports matches, cooking meals, and washing children or their clothes. Chronic pain also reduced the types of physical tasks participants could perform at work, or prevented them from working at all, limiting their ability to provide financial support for dependents. Participants described not being able to perform work-related tasks properly due to chronic pain and needing to adapt their task performance or, in some cases, take leave or resign from their job. Job losses occurred in rare instances, when employers felt participants were no longer fulfilling required duties or excess leave was being taken to manage pain or seek healthcare. Participants described how, when possible, they forced themselves to work despite their pain to financially support younger children:

> *I do my responsibilities as a mother even though I have got the pains. If I need to go to work, I do go to work even if I’ve got the pains because I know that I won’t be able to buy groceries or to buy food for my children if I do not go to work, so even if I’ve got pains, I’ll go to work. I need to go to work. (female in 40s, living with children)*

Some tried to ignore their pain and continue with these necessary tasks, whereas others had to limit their provision of tangible support to their children, which was felt to affect the quality of their relationships with their children or result in their children feeling distressed:

> *I used to play with my kids.. but now after I have the pains, I’m not able to play with them all the time….. they didn’t understand what was happening,… they were saying “Haibo!” [an expression of surprise and disapproval] I don’t want to play at all because of the pains. (female in 20s, living with partner and children)*

Some participants described how their children also became distressed when seeing them in pain or being cared for by others, which in turn caused the participants emotional distress. Others described how older children responded by providing tangible support, taking over the cooking or washing or getting a job to provide financial support. However, some parents preferred not to rely on their children in this way because they perceived task assistance as falling outside a child’s role, made them feel like a burden or too dependent:

> *You know I don’t get happy or satisfied when someone is doing something for me. I just wish I can do it on my own, but I can’t. And even with my children, I don’t get satisfied when they’re doing chores in my house (female in 50s, living with family and children)*

Participants also described how chronic pain influenced their ability to perform domestic and sexual tasks specific to that of being an intimate partner, and how partners had to take over their domestic tasks. Although some described adapting domestic tasks to make them easier or forcing themselves to do them, others described how they often felt too fatigued or in pain, especially in instances where the participant was employed, because work tasks were prioritised rather than domestic chores. At times, this adjustment could cause conflict with partners and complaints:

> *You find that the house is not properly cleaned because I’ve got the pains. Like on Thursday you know, my partner had to make supper. When he was back, he said that, “Haibo, you didn’t do the dishes”. And I told him that I can’t do anything. So, he was complaining that the house is still dirty, the dishes have not been washed, and he didn’t carry any food with to work on that day (female in 30s, living with partner)*

*C*hronic pain also made some participants disinterested in sex, limited how they engaged in sex, or unexpectedly interrupted sex. This further complicated some intimate relationships that were already complicated by participants’ HIV status (i.e. non-disclosure of status, negotiation of condom use, reduced sexual drive perceived to be due to ART use). Although some partners attributed these limitations to pain, others interpreted reluctance to have sex with suspicion, and some participants expressed concerns about how their partner interpreted their reduced sexual activity:

> *There are times where I feel like, okay, and I am supposed to do something [sexual]. Then you find that I’ve got this problem now, I’ve got the pains, I can’t do anything. The thing is maybe if you’ve got someone, and she might not understand you and think that you are you are lazy now, you don’t want to have sex. (male in 40s, living alone)*

Roles external to the household were also described as being affected. For example, participants explained how they could no longer engage in community activities such as sports, helping build friends’ houses, carrying heavy things, or fixing appliances. Some ensured that where they could no longer provide tangible support, they continued to provide companionship and emotional support by visiting their friends and attending community events:

> *Sometimes there is something that is happening in my neighbourhood, maybe they’ve got a traditional work or maybe a funeral or something, I’ll go there, but I’m not able to assist them… I’ll just sit there… I just support them emotionally. (female in 50s, staying alone)*

##### c. Interpersonal relationships can cause emotional distress and pain

Although, as described above, social network members play a crucial role in supporting participants in managing their chronic pain and emotional distress, interpersonal relationships with children, partners and siblings were also a primary source of emotional distress in some cases. Some participants attributed their chronic pain to these interpersonal challenges, specifically chronic headaches, muscle tension in their neck, shoulder and/or back, or severe pain in their sides. Some participants who had lost children (from a motor vehicle accident, fatal assaults and HIV), described their traumatic loss as a sustained source of significant distress:

> *The other thing that is stressing me a lot is the death of my daughter…She had a [detail of unanticipated death], and that is something that will come to my mind most of the time and it made me unhappy…So, that is also causing me to have the pains, that back pain, and you’ll will find that when I think about her, I will have these signs of pain. (female in 50s, living with partner and children)*

Others expressed that their pain was exacerbated or caused by conflict and worrying about their children’s errant behaviours such as smoking, staying out late at night, and not listening:

> *She [my daughter] is stressing me a lot, I will have the pain, like, if I’m talking too much… she will go out and come home very late… I have to speak with her and that was stressing me you know and when I’ve got that stress it will go to my back then I will have pains you know. (female in 40s, living with children)*

Relational challenges with intimate partners caused significant emotional distress and reduced the amount of tangible and emotional social support that could be provided in the relationship. Some participants also described having partners who abused substances, were emotionally unresponsive to their needs, or did not financially support their family or children. One participant also said that she started experiencing her chronic pain when she found out that her partner had married another woman:

> *I was stressing a lot [last year when I started feeling this pain]… I saw a picture of him with another girl and he told me that they got married… now he doesn’t even support the child… He doesn’t support me. (female in 30s, living with family and children)*

A perceived lack of support and open communication from siblings was another source of emotional distress. A participant described moving in with her sister as a form of financial support, but conflict with her sister due to this living arrangement was perceived as the underlying cause for her chronic pain:

> *The most pain that I normally have is the headache, sometimes I will think that I’ve got high blood [pressure], so I went for high blood [pressure] test but I didn’t have it. So, I thought… maybe it’s because I’m stressing a lot, maybe it’s me because I’m so used to having my own space, and now I have to have people that I’m sharing my space with… so, I think stress that is causing this. (female in 40s, living with children and extended family)*

Participants also expressed that their existing chronic pain, and/or other types of pain, were exacerbated or brought on during periods of emotional distress about interpersonal relationships. The result was more intense pain, other locations of pain, or greater difficulties in managing their pain, for example as emotional distress interfered with sleep, as described by a participant experiencing distress over her child’s behaviour below:

> *It is only this thing about my child, I don’t have anything else… The neck and my headache will be intense, will be worse when I’m stressing [about her]…the back and the neck becomes too much… So, if maybe she has stressed me and I will tell myself, I’m not going to speak for days, let me just relax… I don’t respond to anything, then the pains will be better. (female in 40s, living with children)*

## Discussion

This exploratory qualitative study focused on the social network structure and informal social support received by people living with chronic pain and HIV, to address pain and emotional distress. Participants sought support from various people spanning nuclear and extended family members, neighbours, colleagues, friends, and acquaintances within other social networks. Participants sought support from a wider range of people for distress than for pain. Although the nature of support crossed emotional, tangible, informational, and companionship support, many participants perceived benefit from merely speaking to someone who was attentive and empathetic. The analysis revealed that participants saw chronic pain as having the potential to compromise their abilities to fulfil their social roles, which could then disrupt their interpersonal relationships with key providers of informal social support for pain, such as their children and intimate partners. This inability to fulfil social roles, together with social withdrawal (a common strategy to cope with pain or pain-related emotional distress), may render informal social support vulnerable to disruption. Previous work has suggested that people with HIV may socially self-isolate in fear that any ‘display’ of suddenly worsening pain could prompt others to speculate about their overall health and indirectly reveal their HIV status (24, 56). In contrast, our participants attributed their social self-isolation to low mood and functional limitations. Interestingly, in times of interpersonal conflict, the close relationships that usually provided meaningful social support could switch to causing emotional distress and exacerbating, or even causing, pain.

Participants described slightly different support-seeking strategies for pain and emotional distress. To choose a member for distress-focused support, participants considered whether the member would be attentive, their proximity to the problem, the nature and source of the distress, and whether they were well positioned to help. The opportunity to disclose distress was prioritised, and participants seemed less concerned about privacy for distress than for pain. Given the diverse and everyday sources of distress reported, it is possible that normalisation of stressors and worry makes it easier for people to disclose emotional distress in a more public environment. For pain-focused support, participants described a more careful, challenging process of deciding who to disclose to, although availability and access was also a key consideration. For pain, participants also considered the social network member’s general behaviour, discretion, the impact of the knowledge of pain on their roles and responsibilities, and the current health of their relationship with the person. This selectivity echoes that reported in another sample of South Africans with chronic pain (32) and may reflect previous negative interpersonal responses to pain disclosure (57) as well as overlapping HIV and chronic pain stigma, which is thought to contribute to social isolation (58). Notably, in our data, members previously chosen for HIV disclosure were also commonly chosen for pain disclosure. People diagnosed with HIV before chronic pain may benefit from this pre-‘vetted’ circle of trusted others with whom they feel safe to discuss their chronic pain.

This study revealed a potential vulnerability arising from informal social support-seeking behaviours and responses for pain. Participants sought support for pain from a smaller social network than for distress, with the support sought for pain often being more practical and hands-on in nature for household members. High intensity tangible support may be unsustainable over time and result in support fatigue in these smaller support networks (10, 59). As in other literature, participants expressed concerns about shifts in roles, especially between parents and children, and between life partners, describing how this could lead to interpersonal distress and conflict (10, 25). Distrust and additional conflict due to role shifts in relationships—especially in intimate relationships—may reduce social support available for people with HIV and pain if these intimate relationships are disrupted. Analysis of the social networks of people with chronic pain in other contexts suggests that larger social networks are associated with reduced levels of interpersonal conflict, and that inter-personal conflict has a stronger exacerbating effect on distress when the conflict occurs in denser support networks (60). In our participants, if the few people chosen for pain support are closely related to one another, this increase in network density with decreasing network size may render the person with pain particularly vulnerable when conflict does arise.

The current findings stand juxtaposed with previous qualitative studies of people with chronic pain or chronic pain and HIV, most of which were conducted in high-income countries (e.g. 25, 61, 62). In our sample, distress arose largely from concerns that were not specific to HIV or chronic pain, such as difficult living circumstances (poverty, risky housing, income vulnerability, employment problems), interpersonal conflict (sometimes linked to material dependence on others), continued grief, and relational worries. These background stressors may be common to others in the same geographical community, but their diverse and unrelenting nature may represent a higher ‘load’ than in previously described samples, rendering the current sample more vulnerable to pain and negative health outcomes (63). That shifts in functional role performance were a trigger for interpersonal conflict in the current study underscores the limited capacity to absorb further resource deficits. In this context, the recursive, adverse influence of interpersonal conflict on both distress and pain has the potential to contribute to a negative spiral of worsening pain, diminishing function, and continued degradation of social support for the person living with chronic pain and HIV (64).

Interestingly, the impact of chronic pain that was revealed in this qualitative study would be missed by conventional measures of the functional interference of chronic pain. Participants described continuing with their daily roles despite considerable pain, adapting tasks and prioritising employment over home tasks, but rarely completely withdrawing from these. This survival-driven stoicism has been described in resource-strapped contexts and may contrast with pain interference in contexts with higher formal support for people with pain disability (32, 65, 66). For this sample, the true impact of chronic pain is observed in its influence on activities that contribute to relationships and overall wellbeing, rather than just on activities that support survival. Participants described withdrawing from valued activities such as caring for, supporting and playing with their children and expressing physical affection. As in previous South African research (4), participants also described the mental and emotional impact of chronic pain that resulted in worry, low mood, stress and irritability that potentially strained relationships. Such consequences of chronic pain are rarely assessed, but may best capture its true impact on the overall health and wellbeing for the person and their networks, including inter-generational transmission of chronic pain and disability (19, 67). In line with current evidence, the daily impact of living with HIV was almost eclipsed by the impact of living with chronic pain; all participants who drew a comparison declared HIV to be far more easily managed than chronic pain (25, 32, 68).

These qualitative findings provide foundational knowledge for developing and testing interventions to either improve or provide social support for people living with HIV and chronic pain and potentially prevent worsening health. For example, individuals who have existing informal social support networks may benefit from skills to maintain the health of these interpersonal relationships. Training people in pain self-management or equipping people with HIV to better navigate emotionally distressing situations (i.e. through training in emotional self-regulation, workplace negotiation, parenting skills, and conflict resolution) may improve coping skills and reduce the demand on their social network for pain and distress. Similarly, encouraging people with HIV to reciprocate care to members of their informal social support networks could also sustain capacity within these support networks (69). None of these possibilities has yet been tested—but there are existing interventions that could be adapted and tested for training these skills. As examples, the ImpACT+ intervention addresses emotional coping and problem-solving among women with HIV and trauma (70), while the Friendship Bench model supports skill-building using a culturally resonant approach with attractive scalability (71). In the setting of the current research, people with HIV attend monthly ‘adherence club’ meetings to sustain ART adherence. These existing meetings provide an alternative opportunity for brief but regular injections of skills training.

In contrast, for individuals who tend to socially self-isolate, more intentional facilitation of social interactions may be beneficial. One approach could be to use the ART adherence clubs to connect these individuals to a group-based intervention. Group interventions for pain management have been tested for people with HIV, including in a remotely delivered format that may feel more socially ‘safe’ and physically accessible, and some have led to sustained engagements and support between participants (72, 73). Using a trained peer to facilitate such groups is an established practice that is supported by our participants’ reports that they prefer to seek support from people they recognise as having skills or experience. The current participants emphasised the importance of confidentiality, attentive listening, and a sense of common struggles; an intervention that emphasises these features may have better uptake—yet another possibility for future testing. With respect to matching an intervention to an individual, other studies suggest that the individual’s usual preferences about social contact (e.g. intro/extraversion) should inform the type of intervention that is suggested (74).

### Limitations

The timing of this study, the sampling strategy, and the context of the current study inform interpretation of our results. The interviews were conducted in the second half of 2022, when unemployment remained 3% higher than just before the COVID-19 pandemic in 2020 (75, 76). Job losses had disproportionately affected Black South Africans and women, particularly those in face-to-face service jobs like some of our participants (77, 78), and job loss was associated with more depressive symptoms (79). It is likely that this situation influenced participants’ perspectives, and the precarious nature of employment and health-related fears may have exacerbated distress in our sample. The biographical data reported in Table 2 should be interpreted with caution, given that they were collected several months before the interview and some circumstances may have changed – in particular, one participant was no longer living with her children at the time of interview.

**Table 2:** Demographic data as reported by interview participants at enrolment to the parent study. *Multiple selections allowed. ^a^Reported living alone at the time of interview. ^b^Reported living with children at the time of interview.

|  | Sex | Age range | Years with known HIV | Employment* | Housing* | Lives with* |
| --- | --- | --- | --- | --- | --- | --- |
| 1 | Female | 40s | 24 | Full-time | Family | Family, children |
| 2 | Female | 40s | 18 | Unemployed | Family | Children |
| 3 | Female | 30s | 16 | Unemployed, seeking | Shack | Family |
| 4 | Female | 30s | 13 | Seeking | Shack | Partner |
| 5 | Female | 40s | 13 | Unemployed, seeking | Shack | Children |
| 6 | Female | 50s | 12 | Unemployed | Shack | Family, children |
| 7 | Female | 20s | 8 | Unemployed | Shack | Partner, children |
| 8 | Female | 30s | 12 | Full-time | Backyard | Alone, children<sup>a</sup> |
| 9 | Female | 50s | 16 | Casual | Family house | Partner, children |
| 10 | Female | 30s | 15 | Casual | Shack | Partner, children |
| 11 | Female | 50s | 19 | Casual | Shack | Alone |
| 12 | Female | 50s | 13 | Casual | Family house | Children |
| 13 | Female | 30s | 10 | Seeking | Family house | Family, children |
| 14 | Female | 40s | 13 | Unemployed | Shack | Family, children<sup>b</sup> |
| 15 | Male | 50s | 6 | Unemployed | Backyard | Alone |
| 16 | Male | 30s | 3 | Full-time | Shack | Partner, children |
| 17 | Male | 40s | 9 | Unemployed, seeking | Shack | Alone |
| 18 | Male | 40s | 5 | Unemployed, seeking | Backyard | Partner |

We drew participants from a larger study of pain and distress that included in-person assessments, such as venepuncture. Participation in the interviews was voluntary, and people reporting no clinical pain were ineligible. This may have restricted the range of experiences represented in the current dataset – e.g. individuals unwilling to undergo venepuncture in the parent study, individuals unwilling to undergo interview, and individuals reporting no clinical pain could not have their views represented here. Participants may have been more aware of symptoms after frequently being questioned about distress and pain. Having a female lead the interviews may have influenced disclosure by the four male participants, although those interview transcripts seemed to be comparably ‘thick’. Indeed, we judged the interviewer’s existing relationship with the participants to be well suited to foster participant comfort and disclosure and likely contributed to the richness of the data obtained during the interviews. Similarly, the need to analyse, interpret, and report the study findings in English and that analysis was undertaken by two English-speaking, non-Xhosa South Africans, may have resulted in some loss of meaning despite the central involvement of the isiXhosa-speaking interviewer in translation and reviewing data interpretations.

## Conclusion

Informal social support networks for pain are smaller and bear a higher load than those for distress, due to more stringent selection of network members. Informal social support is particularly vulnerable to support fatigue or interpersonal conflict when the person with pain is unable to fulfil their functional social roles, or when they use social isolation to cope with pain and pain-related distress. Conflict in interpersonal relationships can cause and increase pain, emphasising the importance of sustaining good social relationships and support. Individual-level interventions to sustain informal social support should target individuals’ abilities to cope with daily stressors, their knowledge of pain self-management strategies to facilitate socialisation and engagement in valued life activities, and skills to reduce support fatigue in their informal support social network.

## Data Availability

The de-identified study transcripts are available for selective sharing,
subject to review of individual data requests, the use of secure computer platforms, formal use agreements, and compliance with the institutional human research ethics policies. Data use is limited to research purposes, on secure computer servers. The principal investigator (VJM) is the contact person for requests to share data. Raw audio-recordings are not available for sharing.

https://osf.io/kbfz9/overview?view_only=812bb52be753456bb059d75e08e5922b

